# Body fat and human cardiovascular ageing

**DOI:** 10.1101/2024.06.27.24309526

**Authors:** Vladimir Losev, Chang Lu, Deva S Senevirathne, Paolo Inglese, Wenjia Bai, Andrew P King, Mit Shah, Antonio de Marvao, Declan P O’Regan

## Abstract

**Objective:** Cardiovascular ageing is a progressive loss of physiological reserve, modified by environmental and genetic risk factors, that contributes to multi-morbidity due to accumulated damage across diverse cell types, tissues and organs. Obesity is implicated in premature ageing but the effect of body fat distribution in humans is unknown. Here we assessed the association between image-derived adiposity phenotypes on cardiovascular age in men and women.

**Methods:** We analysed data from 21,241 participants in UK Biobank. Machine learning was used to predict cardiovascular age from 126 image-derived traits of vascular function, cardiac motion and myocardial fibrosis. An age-delta was calculated as the difference between predicted age and chronological age. The volume and distribution of body fat was assessed from whole body imaging. The association between adiposity phenotypes and cardiovascular age-delta was assessed using multivariable linear regression with age and sex as co-covariates, reporting β coefficients with 95% confidence intervals (CI). Two-sample Mendelian randomisation was used to assess causal associations.

**Results:** Visceral adipose tissue volume (β = 0.656, [95% CI, 0.537 - 0.755], *P* < 0.0001), muscle adipose tissue infiltration (β = 0.183, [95% CI, 0.122 - 0.244], *P* = 0.0003), and liver fat fraction (β = 1.066, [95% CI 0.835 - 1.298], *P* < 0.0001) were the strongest predictors of increased cardiovascular age-delta for both sexes. Abdominal subcutaneous adipose tissue volume (β = 0.688, [95% CI, 0.64 - 1.325], *P* < 0.0001) was associated with increased age-delta only in males. Genetically-predicted gluteo-femoral fat showed an association with decreased age-delta.

**Conclusion:** This work demonstrates the contribution of sex-dependent patterns of visceral adipose tissue and subcutaneous fat distribution to biological cardiovascular ageing in middle aged adults. This highlights the potential for strategies to attenuate ageing through modifying adipose tissue function.

## Introduction

Obesity is a chronic complex condition characterised by excessive fat deposits that are detrimental to health. Globally 43% of adults, affecting men and women in equal proportions, are overweight with a rising prevalence.^1^ It is a multifactorial and heterogeneous disease related to obesogenic environments, psycho-social influences and genetic risk factors. Individuals with similar body mass index (BMI) may have distinct metabolic and cardiovascular disease (CVD) risk profiles, therefore susceptibility to obesity-related cardiovascular complications is not mediated solely by overall body mass but may be strongly influenced by variation in fat distribution.^2^ For instance, visceral fat confers a higher risk of adverse outcomes,^3^ promotes systemic and vascular inflammation,^4,5^ and is independently linked with dysmetabolic profiles.^6^

Obesity, as a systemic syndrome of adipose tissue dysfunction, can be considered a pro-inflammatory process of accelerated cardiovascular ageing with which it shares common genetic, epigenetic, immunological and metabolic mechanisms.^7,8^ Recently, imaging and physiological parameters have been used to assess environmental and genetic risk factors for accelerated biological ageing which point to complex inter-organ ageing networks that are regulated by genes involved in inflammation, tissue elasticity and pro-fibrotic pathways.^9,10^ Relatively less is known about drivers of sexual dimorphism in cardiovascular ageing, although cellular and molecular mechanisms of ageing may be better maintained in women until the menopause.^11^ Sex hormones modify key aspects of nutrient sensing, metabolism and fat depot regulation - with men tending to have more visceral fat, whereas women have greater fat deposition in the lower body.^12^ The role of sex-specific fat distribution on accelerated cardiovascular ageing is not known, but could be a key modifiable mediator of obesity-related risk.

In this study, we use machine learning techniques to predict biological age from multiple image-derived cardiovascular traits that assess the structure, function and tissue characteristics of the heart and circulation. We then express how an individual’s cardiovascular system has aged relative to a normative population using an “age-delta”. Through assessing body fat distributions with whole body image phenotyping, as well as circulating biomarkers and sex hormones, we aimed to understand the relationship between fat distribution and cardiovascular ageing in over 20,000 middle aged men and women.

## Methods

### Study overview

UK Biobank comprises approximately 500,000 community-dwelling participants aged 40–69 years who were recruited across the United Kingdom between 2006 and 2010.^13^ All participants provided written informed consent for participation in the study, which was approved by the National Research Ethics Service (11/NW/0382). Our study was conducted under terms of access approval number 40616.

An outline of the methods is shown in Figure 1. We used a pre-trained model to predict cardiovascular age from 126 image-derived traits of vascular function, cardiac motion and myocardial fibrosis. Cardiovascular age-delta was calculated as the difference between predicted age and chronological age.^9^ We combined data from body imaging to assess phenotypes of fat volume and distribution including visceral adipose tissue, abdominal subcutaneous adipose tissue, muscle adipose tissue infiltration, liver proton density fat fraction, total abdominal adipose tissue, android adipose tissue mass and gynoid adipose tissue mass, as well as total trunk and whole body fat mass.

**Figure 1.**
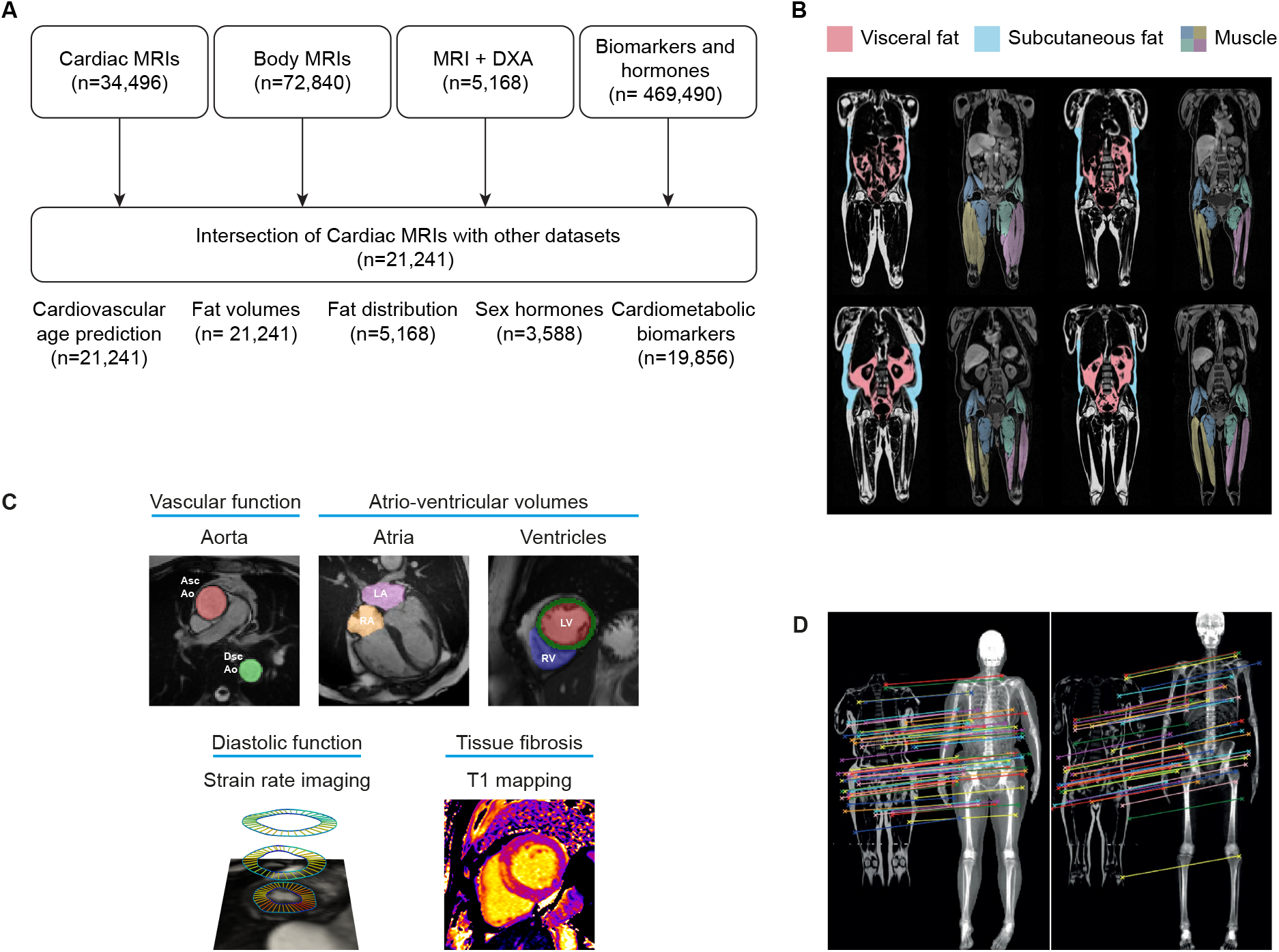
Analysis of fat phenotypes and cardiovascular aging. **A** Flowchart of analyses performed in UK Biobank participants. **B** Fat phenotyping was performed by segmentation of whole body MRI into visceral, subcutaneous and muscle compartments (credit: AMRA Medical). **C** Phenotypes derived from cardiac MRI were used for age prediction. These included automated time-resolved segmentations of the aorta and cardiac chambers, as well as strain rate analysis and T1 mapping. **D** Integration of MRI and DXA enabled regional body composition analysis. (credit: Rhydian Windsor,^24^ under Creative Commons Attribution License CC BY 4.0). MRI, magnetic resonance imaging; DXA, dual X-ray absorptiometry; Asc Ao, ascending aorta; Dsc Ao, descending aorta.

We used multivariable linear regression modelling stratified by sex to assess the association of fat phenotypes with cardiovascular ageing. Additionaly, the relationship between BMI categories and fat phenotypes by sex was assessed.

### Cardiac image acquisition

A standardised cardiac magnetic resonance (CMR) imaging protocol was followed to acquire two-dimensional, retrospectively-gated cine imaging on a Siemens MAGNETOM Aera 1.5-T scanner (Siemens Healthineers, Erlangen, Germany).^14^ Short-axis cine imaging comprised a contiguous stack of images from the left ventricular base to apex, and long axis cine imaging was performed in the two and four chamber planes. Cine sequences consisted of 50 cardiac phases with an acquired temporal resolution of 31 ms.^14^ Transverse cine imaging of the ascending and descending thoracic aorta was also performed. Native T1 mapping within a single breath hold was performed at mid-ventricular level using a shortened modified Look-Locker inversion recovery (ShMOLLI) sequence. Imaging phenotypes all underwent quality control prior to use in analysis.^15^

### Cardiac image analysis

Automated segmentation of the short-axis and long-axis cine images in UK Biobank was performed using fully convolutional networks.^16^ Volumes (end-diastolic, end-systolic, and stroke volume) and ejection fraction were determined for both ventricles. Myocardial volumes were used to compute left ventricular myocardial mass assuming a density of 1.05 g.ml ^−1^. Atrial volumes were calculated using biplane area–length. Central vascular function was assessed by measuring aortic distensibility from central blood pressure estimates and dynamic aortic imaging.^17^ The aorta was segmented on the cine images with a spatio-temporal neural network,^18^ from which maximum and minimum cross-sectional areas were derived. Distensibility was calculated using central blood pressure estimates obtained using peripheral pulse-wave analysis (Vicorder, Wuerzburg, Germany).^17^

Diastolic function, which is a key feature of the ageing heart, was assessed using motion analysis to derive end diastolic strain rates.^16^ Non-rigid image registration between successive frames enabled motion tracking on greyscale images.^19^ Registration errors were minimised by tracking motion in both backwards and forwards directions from end-diastole, resulting in an averaged displacement field,^17^ which was then used to warp segmentations from end-diastole to successive adjacent frames. Circumferential (*E*_cc_) and radial (*E*_rr_) strains were calculated using short axis cines. Longitudinal (*E*_ll_) strain was calculated from long-axis four-chamber motion tracking measured at basal, mid-ventricular, and apical levels. Segmental and global peak strains were then calculated. Strain rate was computed as the first derivative of strain and thereafter peak early diastolic strain rate in radial (PDSR_rr_) and longitudinal (PDSR_ll_) directions calculated. We assessed diffuse myocardial fibrosis, an early feature of natural ageing,^20^ using native T1 mapping of the interventricular septum.^21^ The ShMOLLI T1 maps were analysed using probabilistic hierarchical segmentation with automated quality control defining a region of interest within the interventricular septum as previously validated.^21^ Blood pool T1 was used as a linear correction of myocardial T1 values.^21,22^

In total, 126 quantitative imaging phenotypes characterising structure, function and tissue characteristics were generated for each participant.

### Cardiovascular age prediction

We used a model to predict cardiovascular age from image-derived phenotypes that was pre-trained on 5,065 healthy individuals, not included in the current study, that were free of cardiac, metabolic or respiratory disease and had a body mass index below 30. The development and performance of the model has been previously reported (Supplementary Methods).^9^ Briefly, we used CatBoost, a machine learning algorithm based on decision trees and gradient boosting, to estimate the age of each participant from a joint analysis of all cardiac image-derived phenotypoes. Similar to brain age modelling, we addressed the correlation between age-delta and chronological age using a linear regression analysis between the initial unadjusted cardiovascular age-delta and chronological age. Subsequently, an offset was determined by multiplying the chronological age by the slope of the regression line and adding the intercept. This offset was then subtracted from the initial unadjusted predicted age to yield the corrected predicted age.^9^

### Body adipose tissue analysis

Abdominal and body adipose tissue assessment was performed on the same 1.5T scanner using a dual-echo Dixon Vibe imaging protocol, enabling a comprehensive evaluation of the entire body from the neck to the knees.^23^ This imaging protocol generated a dataset with water and fat components separated, facilitating the analysis of body fat composition. In brief, 6 overlapping sections were acquired that underwent calibration, stacking, fusion, and segmentation. Body composition analyses were carried out using the AMRA Profiler™ (AMRA AB, Linköping, Sweden). Values for android and gynoid adipose tissue mass were acquired from over 20,000 subjects UK Biobank dataset with multi sequence magnetic resonance and dual-energy X-ray absorptiometry (DXA) scans, using self-supervised, multi-modal alignment for whole body medical imaging, and transferring segmentation maps from DXA to MRI scans achieving high accuracy in matching different-modality scans without requiring ground-truth magnetic resonance examples .^24^

We also utilised datasets for liver proton density fat fraction employing the gradient echo protocol from the UK Biobank, specifically the LMS (Liver MultiScan) Dixon method. The LMS Dixon proton density fat fraction captures images during a single breath-hold, reducing motion artifacts and ensuring consistent measurements. Analysis involves three 15 mm circular regions of interest in the liver parenchyma, calculating proton density fat fraction, liver iron concentration, and iron-corrected T1 (cT1). Proton density fat fraction, a reliable measure of liver fat, is determined using water-fat separation masks, with values over 5% indicating fatty liver disease.^25^

Standing height without shoes was assessed using a Seca (Hamburg, Germany) 202 height measure. Weight, without shoes or outer clothing, was assessed with a Tanita (Tokyo, Japan) body composition analyser. Each BMI was calculated as weight in kilograms divided by height in metres^2^.

### Circulating bio-markers

We assessed 9 cardiometabolic and endocrine circulating biomarkers (triglycerides, direct low-density lipoprotein (direct LDL) cholesterol, high density lipoprotein (HDL) cholesterol, cholesterol (total), apolipoprotein A and B, sex hormone binding globulin (SHBG), oestradiol (E2), testosterone (free form)) as potential modifiers of cardiovascular ageing. The collection of these biomarkers involved standardised venous blood sampling followed by serum separation and storage at −80°C to ensure stability. The samples were then analysed using validated biochemical assays to quantify levels of each biomarker.^26^

### Mendelian randomisation

Mendelian randomisation (MR) was performed to investigate the potential causality of adipose tissue volumes on cardiovascular age-delta. We used large-scale genome wide association studies of image-derived visceral, abdominal subcutaneous, and gluteofemoral adipose tissue volumes after adjustment with BMI and height,^27^ as well as of the cardiovascular age-delta.^9^

We used the R package TwoSampleMR to perform the analysis.^28^ Independent genetic instruments were selected at the conventional genome wide significance threshold of *P* < 5 × 10^−8^, using the 1000G genomes European population as reference for linkage disequilibrium (LD) with an r^2^ threshold of 0.001. Exposure and outcome statistics were harmonised to allow consistency of alleles for MR analysis. Palindromic single nucleotide polymorphisms (SNPs) with intermediate allele frequencies were removed before MR. For each analysis, we performed MR with 5 methods,^28^ including the inverse variance-weighted method that assumes all genetic variants are valid IVs, the MR Egger regression as a check for horizontal pleiotropy, the weighted median method, which is additionally robust in the presence of outliers, the simple mode and weighted mode methods which clusters SNPs into groups based on similarity of causal effects.

### Statistics

Analysis was performed in R (version 4.2.3) and Python. Variables were expressed as percentages and frequencies if categorical, mean ± standard deviation (SD) if continuous and normal, and median ± inter-quartile range (IQR) if continuous and non-normal. Comparison of independent groups was performed with a two sample *t* test. Multivariable linear regression was used to estimate the association between age-delta (as the dependent variable), and adipoisity phenotypes and cardiometabolic biomarkers (as independent variables) using age, age^2^, and sex as covariates. Additionally, the model was adjusted for height^2^ to account for variation in stature (Supplementary Methods).^29^ Standardised β coefficients are reported for continuous predictors, each with 95% confidence intervals (CI). Fat mass was categorised using centile ranges of each BMI group and then compared with alluvial plots (see Supplementary Methods and Supplementary Figure 1). We applied the Benjamini-Hochberg procedure to control the false discovery rate. Statistical significance was considered at *P* ≤ 0.05 (two-sided).

## Results

### Sex-dependent associations of body fat with chronological age

In total 21,241 participants were included in the analysis (Table 1). Females had more abdominal subcutaneous fat (8.3 ± 3.5 L vs 6.0 ± 2.5 L, *P* < 0.0001), muscle adipose tissue infiltration (7.9 ± 1.9 % vs 6.9 ± 1.7 %, *P* < 0.0001) and gynoid fat (4.8± 1.6 kg vs 3.6 ± 1.2 kg, *P* =0.0001), whereas males predominantly had greater volumes of visceral fat (5.1 ± 2.3 L vs 2.8 ± 1.5 L, *P* < 0.0001), android fat (2.8 ± 1.2 kg vs 2.3 ± 1.2 kg, *P* = 0.0002), and whole body fat mass (23.9 ± 8.6 kg vs 20.1 ± 7.0 kg, *P* < 0.0001) (Figure 2). Muscle adipose tissue infiltration increased by an average of 11.7% per decade in females and 9.8% in males. Visceral fat increased more with age in males, rising by an average of 8.2% each decade compared to 5.3% in females. Abdominal subcutaneous fat showed a modest decrease of 5.4% in females and 4.3% in males per decade (Figure 3). Adipose phenotype distribution by ancestry is shown in Supplementary Figure 2 and Supplementary Table 1.

**Table 1.** Study population characteristics. Baseline cardiac MRI and adipose tissue parameters by sex.

|  | Female (n = 10558)<br>Mean ± SD, or n (%) | Male (n = 10683)<br>Mean ± SD, or n (%) | P value |
| --- | --- | --- | --- |
| **Baseline characteristics** |  |  |  |
| Age at MRI (years) | 62.5 ± 7.3 | 63.9 ± 7.6 | <0.0001 |
| White ancestry (n/%) | 9690/91.7 | 9822/91.9 | 0.68 |
| Black ancestry (n/%) | 181/1.7 | 226/2.1 | 0.037 |
| Asian ancestry (n/%) | 386/3.7 | 296/2.8 | 0.0009 |
| Mixed ancestry (n/%) | 301/2.9 | 339/3.2 | 0.18 |
| Body mass index | 26.6 ± 4.9 | 27.3 ± 3.9 | <0.0001 |
| Systolic blood pressure (mmHg) | 131 ± 17 | 139 ± 18 | <0.0001 |
| Diastolic blood pressure (mm Hg) | 77 ± 8 | 82 ± 8 | <0.0001 |
| Pulse rate (bpm) | 75 ± 7 | 71 ± 8 | <0.0001 |
| Diabetes mellitus (n/%) | 745/7.1 | 1088/10.2 | <0.0001 |
| Hypercholesterolaemia (n/%) | 975/9.2 | 1749/16.3 | <0.0001 |
| Hypertension (n/%) | 2129/20.2 | 3082/28.8 | <0.0001 |
| Coronary artery disease (n/%) | 383/3.6 | 1162/10.8 | <0.0001 |
| Obesity (n/%) | 2491/23.6 | 2826/26.5 | <0.0001 |
| Current smoker (n/%) | 383/3.6 | 508/4.8 | <0.0001 |
| Current alcohol consumption (n/%) | 338/3.2 | 483/4.5 | <0.0001 |
| **Cardiac parameters from CMR** |  |  |  |
| Left ventricular ejection fraction (LVEF) (%) | 60.7 ± 5.7 | 58.2 ± 6.2 | <0.0001 |
| Left ventricular end-diastolic volume (LVEDV) (ml) | 133.6 ± 26.5 | 164.3 ± 33.6 | <0.0001 |
| Left ventricular end-systolic volume (LVESV) (ml) | 52.8 ± 14.9 | 69.2 ± 20.2 | <0.0001 |
| Left ventricular stroke volume (LVSV) (ml) | 80.8 ± 16.0 | 95.1 ± 19.7 | <0.0001 |
| Left ventricular cardiac output (LVCO) (ml) | 5.1 ± 1.1 | 5.9 ± 1.3 | <0.0001 |
| Left ventricle mass (LVM) (g) | 74.5 ± 16.1 | 100.0 ± 20.3 | <0.0001 |
| Right ventricular ejection fraction (RVEF) (%) | 58.8 ± 5.7 | 55.7 ± 6.1 | <0.0001 |
| Right ventricular end-diastolic volume (RVEDV) (ml) | 138.9 ± 29.3 | 175.6 ± 35.5 | <0.0001 |
| Right ventricular end-systolic volume (RVESV) (ml) | 57.6 ± 16.3 | 78.2 ± 20.6 | <0.0001 |
| Right ventricular stroke volume (RVSV) (ml) | 81.3 ± 17.0 | 97.3 ± 20.5 | <0.0001 |
| **Adiposity parameters from MRI** |  |  |  |
| Visceral adipose tissue (VAT) (L) | 2.76 ± 1.5 | 5.1 ± 2.3 | <0.0001 |
| Abdominal subcutaneous adipose tissue (ASAT) (L) | 8.3 ± 3.5 | 6.0 ± 2.5 | <0.0001 |
| Muscle adipose tissue infiltration (MATI) (%) | 7.9 ± 1.9 | 6.9 ± 1.7 | <0.0001 |
| Liver proton density fat fraction (PDFF) ( %) | 4.8 ± 4.3 | 4.6 ± 4.4 | 0.60 |
| Whole body fat mass (WBFM) (kg) | 20.1 ± 7.0 | 23.9 ± 8.6 | <0.0001 |
| Total trunk fat mass (TTFM) (kg) | 14.4 ± 5.0 | 12.0 ± 4.8 | <0.0001 |
| Gynoid fat mass (kg) | 4.8±1.6 | 3.6±1.2 | 0.0001 |
| Android fat mass (kg) | 2.3±1.2 | 2.8±1.2 | 0.0002 |

**Figure 2.**
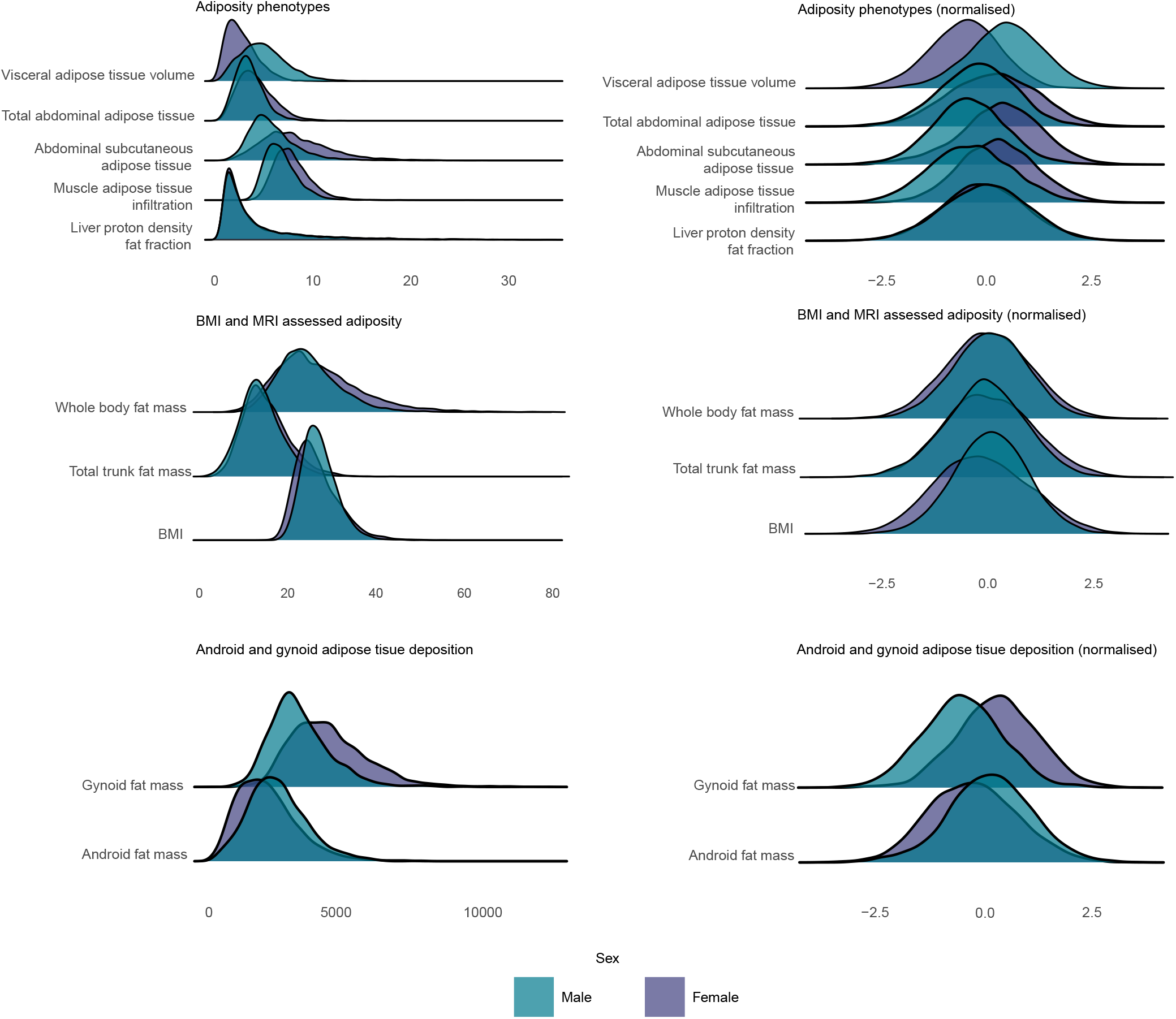
Distribution of fat phenotypes and association with cardiovascular age-delta. Ridge plots summarising the distribution densities of adiposity phenotypes. Unadjusted and normalised values shown. Body mass index, BMI; Magnetic resonance imaging, MRI; Adiposity phenotypes n= 21,241; BMI and MRI assessed adiposity n= 21,241; Android and gynoid adipose tissue deposition n = 5,168.

**Figure 3.**
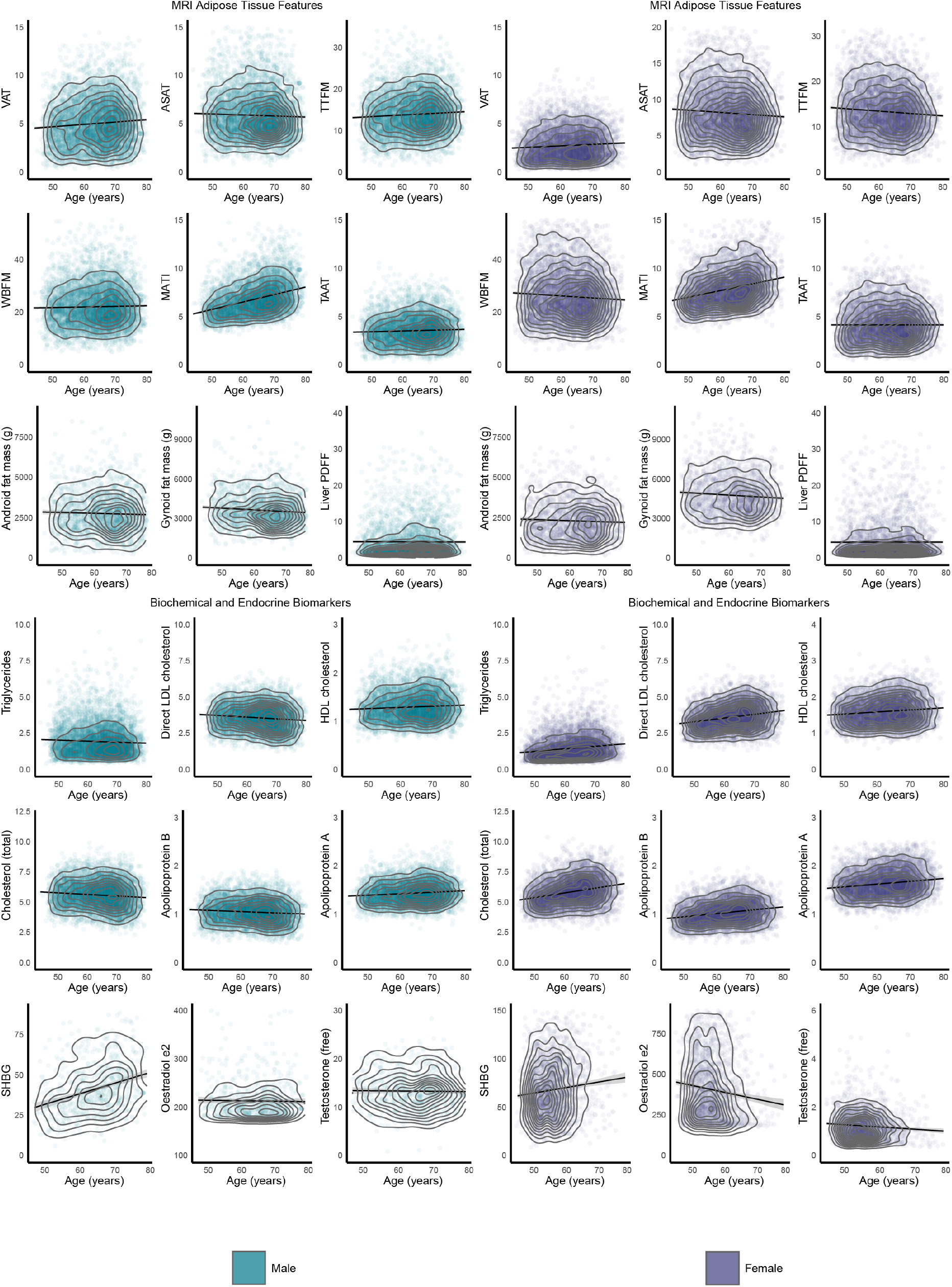
Adiposity and biomarker associations with chronological age. A selection of representative phenotypes grouped by MRI-derived adipose features (n=21,241), android and gynoid fat mass (n=5,168), circulating biomarkers (n=19,856) and hormones (n=3,588) are shown with their relationship to chronological age at the time of imaging (ages jittered, density contours, point colours represent coefficient of determination (R^2^). VAT, visceral adipose tissue; ASAT, abdominal subcutaneous adipose tissue; TTFM total trunk fat mass; WBFM, whole body fat mass; MATI, muscle adipose tissue infiltration; TAAT, total abdominal adipose tissue; PDFF, proton density fat fraction (of the liver); LDL, low density lipoprotein; HDL, high density lipoprotein; SHBG, sex hormone-binding globulin.

### Association of cardiovascular age-delta and body fat phenotypes

Visceral adipose tissue (β = 0.656, [95% CI, 0.537 - 0.775], *P* <0.0001), muscle adipose tissue infiltration (β = 0.448, [95% CI, 0.112 - 0.224], *P* = 0.0005), liver fat fraction (β = 1.066, [95% CI, 0.835 - 1.298], *P* < 0.0001), and total abdominal adipose tissue (β = 0.615, [95% CI, 0.499 - 0.732], *P* < 0.0001) were associated with adverse changes in cardiovascular age-delta for both sexes. For males, an increased age-delta was associated with android (β = 0.983, [95% CI, 0.64 - 1.326], *P* < 0.0001) and gynoid fat (β = 0.688, [95% CI, 0.33 - 1.046], *P* = 0.0066) (Figure 4A). Conversely, in females, gynoid fat (β = −0.499, [95% CI, −0.85 - −0.149], *P* = 0.0003), total trunk fat mass (β = −0.403, [95% CI, −0.821 - −0.14], *P* = 0.0061), and whole-body fat mass (β = −0.389, [95% CI, −0.732 - −0.254], *P* = 0.0043) were associated with beneficial changes in cardiovascular age-delta (Table 2). BMI was a weaker predictor of age-delta than body fat in females (β = −0.85, [95% CI, −0.115 - −0.055], *P* = 0.0092) and males (β = 0.063, [95% CI, 0.026 - 0.1], *P* = 0.0813).

**Table 2.**
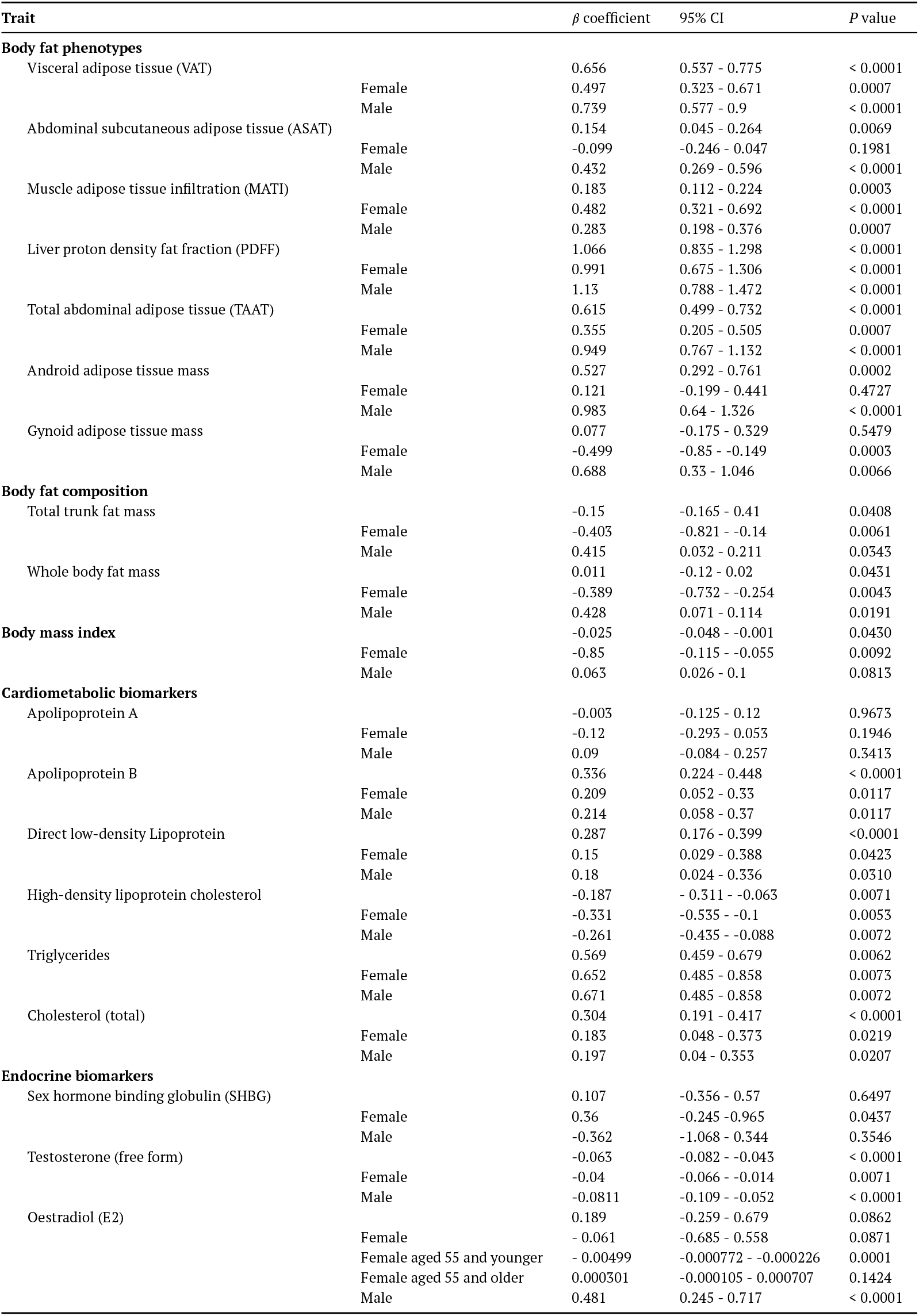
Association of adiposity phenotypes, cardiometabolic and endocrine biomarkers with cardiovascular age-delta. Sex stratified β coefficients, 95% confidence intervals and *P* values for each predictor with age-delta as the dependent variable in a linear regression model with age and age^2^ as covariates.

**Figure 4.**
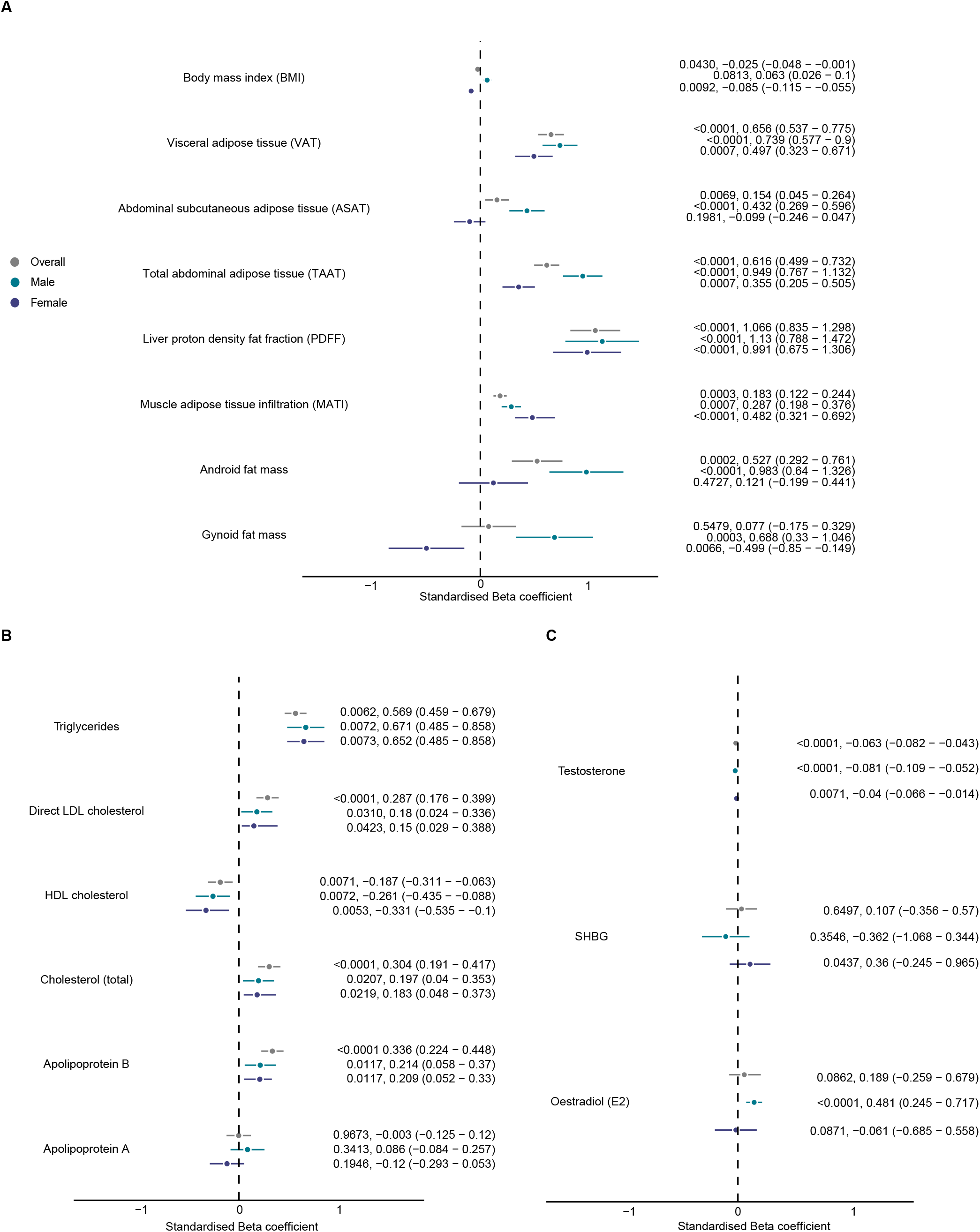
Adiposity phenotype, cardiometabolic and endocrine associations with cardiovascular age-delta. **A** Linear regression analysis of quantitative adipose tissue traits (n=21,241, of which 5,168 had android and gynoid fat mass values) with cardiovascular age-delta as the dependent variable. *P* values, standardised beta-coefficient point estimates, and 95% confidence intervals shown stratified by sex. Linear regression analysis of **B** circulating lipids (n=19,856) and **C** sex hormones (n=3,588) with cardiovascular age-delta as the dependent variable. *P* values, standardised beta-coefficient point estimates, and 95% confidence intervals shown stratified by sex. LDL, low density lipoprotein; HDL, high density lipoprotein; SHBG, sex hormone-binding globulin.

The relationship between fat distribution and cardiovascular age-delta remained mainly independent of height^2^. Visceral adipose tissue (β = 0.432, [95% CI, 0.296 - 0.567], *P* <0.0001), liver fat fraction (β = 0.207, [95% CI, 0.154 - 0.261], *P* <0.0001), muscle adipose tissue infiltration (β = 0.21, [95% CI, 0.055 - 0.364], *P* = 0.0078) and total abdominal adipose tissue (β = 0.326, [95% CI, 0.165 - 0.487], *P* < 0.0001) remained the strongest predictors for increased age-delta in the overall cohort. However in females muscle adipose tissue infiltration (β = 0.08, [95% CI, −0.116 - 0.277], *P* = 0.4235), total trunk (β = 0.003, [95% CI, −0.068 - 0.075], *P* = 0.9276) and whole body fat mass (β = −0.004, [95% CI, −0.042 - 0.034], *P* = 0.8255) became insignificant, while gynoid fat mass (β = −0.499, [95% CI, −0.85 - −0.149], *P* = 0.0052) showed a stronger negative association with age-delta (Supplementary Figure 3).

### Mendelian randomisation analysis

Using 17, 14, and 25 genetically independent instruments, we tested the causal association of image-derived fat phenotypes with cardiovascular age-delta (Supplementary Table 2). We observed genetic associations of gluteofemoral adipose tissue to cardiovascular age-delta (β = −0.96, [95% CI, −0.39 - −1.52], *P* = 8.7 × 10^−4^) from inverse variance-weighted two-sample MR, suggesting a potentially protective role against increasing cardiovascular age-delta. Two other fat traits, visceral and abdominal subcutaneous adipose tissue, did not have significant association with age-delta but had the opposite effect directions to gluteofemoral adipose tissue (Supplementary Table 3 and Supplementary Figure 4).

### Relationship between circulating biomarkers and cardiovascular age-delta

We found significant associations between increased cardiovascular age-delta and apolipoprotein B (β = 0.336, [95% CI, 0.224 - 0.448], *P* < 0.0001), total cholesterol (β = 0.304, [95% CI, 0.191 - 0.417], *P* < 0.0001), and direct LDL cholesterol (β = 0.287, [95% CI, 0.176 - 0.399], *P* < 0.0001) while HDL cholesterol showed a decrease in cardiovascular age-delta (β = −0.187, [95% CI, −0.311 - −0.0063], *P* =0.0071) across both sexes. Notably, no significant changes between sexes were observed (Figure 4B).

Oestradiol (E2) was associated with increased cardiovascular age-delta in males (β = 0.481, [95% CI, 0.245 - 0.717], *P* < 0.0001) while in all-age females it did not show a significant association (β = −0.061, [95% CI, −0.685 - −0.558], *P* = 0.0871). However, in females aged ≤ 55 years, oestradiol was associated with a weak effect on decreased cardiovascular age-delta (β = −0.000499, [95% CI, −0.000772 to −0.000226], *P* = 0.0001) (Table 2). An association between sex hormone-binding globulin (SHBG) and increased age-delta was seen in females (β = 0.360, [95% CI, −0.245 - 0.965], *P* = 0.0437). Free testosterone was associated with a decrease in cardiovascular age-delta in both sexes (males: β = −0.081, [95% CI, −0.109 - −0.052], *P* < 0.0001; females: β = −0.04, [95% CI, −0.066 - −0.014], *P* = 0.0071) (Figure 4C).

### Comparison of body mass index with fat mass

Categorising body composition by the same centile ranges as BMI-defined normal, overweight and obese categories we showed that 31% (n=1141/3681) of overweight female participants fell into the “normal” range for whole body fat mass. Among overweight male participants, 11% (n=537/4882) were reclassified to “normal” whole body fat mass, and 23% (n=1123) into the “obese” range (Figure 5).

**Figure 5.**
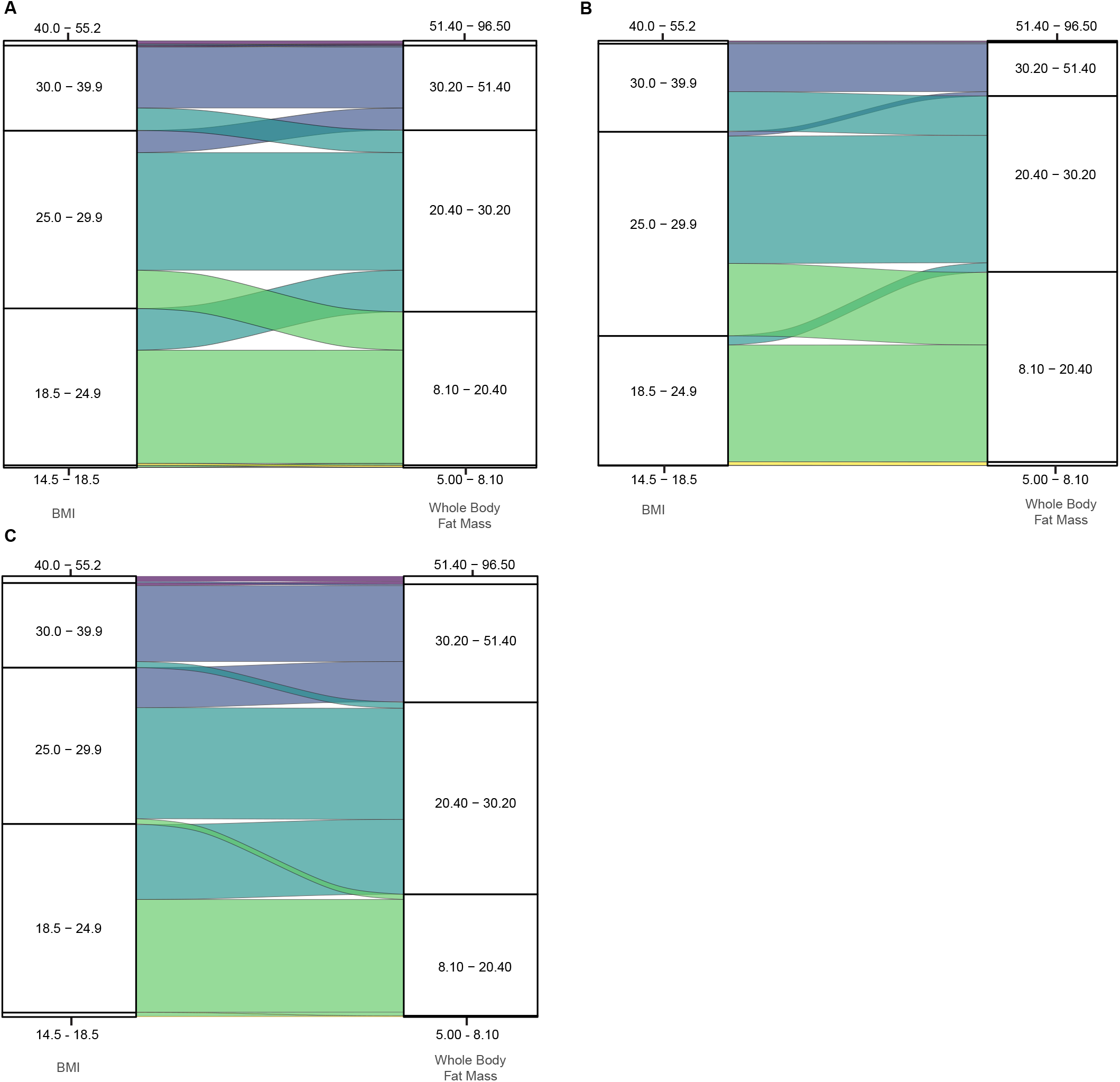
Reclassification of body mass index groups by whole body fat mass. Series of alluvial plots that show the redistribution of participants in each body mass index (BMI) group to equivalent centile ranges of whole body fat mass. **A** Overall population (n=21,241), **B** females (n=10,558), and **C** males (n=10,683).

## Discussion

Obesity is a leading cause of CVD independent of other risk factors,^30^ and in animal models causes a state of accelerated biological ageing of the heart and circulation through up-regulation of pro-fibrotic and inflammatory factors.^31,32^ Here we show that fat distribution may underlie premature ageing of the human cardiovascular system and that phenotypic differences between sexes modify these processes. This work demonstrates the key role played by visceral adipose tissue and the distribution of subcutaneous fat in regulating biological age and highlights the potential value for novel therapies intended to extend health span through modifying adipose tissue function.

Adipose tissue is a metabolically active distributed organ system composed of a variety of cell types that store energy and also contribute to the regulation of diverse biological functions through secretion of cytokines, chemokines, and hormones.^33^ Adipose tissue is morphologically heterogeneous with white, beige and brown fat possessing increasing thermogenic adipocytes.^34^ Visceral and subcutaneous white adipose tissue depots are also developmentally distinct,^35^ with visceral adiposity associated with insulin resistance, local and systemic inflammation, and dyslipidemia.^36^ We showed that absolute visceral fat volume in women is 54% of that seen in men while subcutaneous fat is 38% higher than men. In middle aged adults, while whole body fat remains relatively stable with age there is a progressive increase in muscle fat infiltration, a small rise in visceral fat volume and a decline in subcutaneous fat in both sexes. Such age-related changes in adipose tissue are thought to involve a redistribution of fat depots and changes in their cellular composition, alongside a functional decline of adipocyte progenitors and accumulation of senescent cells.^37^ In animal models, widespread activation of immune cells is especially pronounced with the earliest signs present in white adipose depots during middle age as part of an asynchronous pattern of inter-organ ageing.^38^

We used non-invasive imaging to provide an estimate of how an individual’s cardiovascular system has aged relative to a normative population and explore the association with fat phenotypes.^9^ We found that BMI was a weak predictor of age-delta in either sex reflecting that accelerated ageing is not predicted by overall body mass. BMI also showed a sex bias in terms of over-representing women with normal fat mass as overweight and *vice versa* for men. Our data showed that visceral adipose tissue, liver fat, and to a lesser extent, muscle fat infiltration all predicted an increased age-delta in both sexes. Visceral adipose tissue promotes abnormal secretion of adipose-derived inflammatory cytokines and bioactive peptides which are thought to promote premture brain ageing,^39,40^ and here we show a potential shared mechanism with accelerated ageing of the heart and circulation that is independent of sex. We also found key sex differences with a gynoid fat distribution appearing protective for ageing in females and a potential causal association of genetically-determined gluteofemoral fat on attenuated cardiovascular ageing. Gluteofemoral fat is negatively correlated with cardiometabolic disease risk factors,^41^ and while the mechanism remains unclear it may be partially mediated by secretion of adiponectin which enhances insulin sensitivity.^41^ However, gynoid fat was only associated with attenuated ageing in women suggesting that hormonal factors that regulate fat distribution could also directly influence ageing.^42^ Oestradiol plays a key role in the biology of ageing across organ systems,^11,43,44^ and we found that it might attenuate cardiovascular ageing in women until the menopause.

Our observation that visceral fat promotes ageing of the heart and circulation in humans provides support for the potential role of emerging treatments that target adipose tissue function to extend health-span. Glucagon-like peptide-1 receptor agonists (GLP-1 RAs), a class of antihyperglycemic drugs, are used to manage type 2 diabetes mellitus, but also have pleiotropic effects on protecting against age-related oxidative stress, cellular senescence and chronic inflammation. They also substantially reduce visceral and liver fat in those with or without diabetes.^45^ Therefore they may reduce both the volume of visceral fat as well as suppress secreted pro-inflammatory mediators of ageing.^46,47^ For age-associated diseases and lifespan, ERK, AMPK and mTORC1 represent critical modifiable pathways.^48^ In animal models of ageing IL11 is progressively up-regulated in liver, skeletal muscle, and fat to stimulate an ERK/AMPK/mTORC1 axis of cellular, tissue and organismal ageing pathologies. Anti-IL11 therapy may reactivate age-repressed metabolic function in adipose tissue and is a potential therapeutic target for extending mammalian healthspan.^49^ Together, this shows the potential for treatments that target “inflamm-ageing” through mechanisms that include reprogramming of adipose tissue function.

Our study has limitations. Older age groups and persons living in less socioeconomically deprived areas are under-represented in UK Biobank.^50^ The population is predominantly European and further work is required in people of diverse ancestries and social groups. People of Asian ancestry in particular may have different sex-dependent distributions of fat compared to other ancestries.^51^ Phenotyping is derived at a single time-point in this cross-sectional study, and we could not assess within-person trajectories nor fully account for differential cohort and periodic effects.

In conclusion, sex-dependent fat phenotypes are related to biological cardiovascular ageing in humans which highlight adipose tissue distribution and function as potential targets for interventions to extend healthy lifespan.

## Author contributions

Conceptualization: D.P.O’R.; Methodology: V.L., P.I.; Formal Analysis: V.L., C.L.; Resources: A.P.K., M.S., A. de M., W.B.; Software: P.I., D.S., V.L.; Writing – Original Draft: V.L., D.P.O’R.; Writing – Review & Editing: (all authors); Funding Acquisition: D.P.O’R.

## Data availability

The scripts for data analysis are publicly available https://github.com/ImperialCollegeLondon/adiposity_aging. Data from UK Biobank is available for approved research.

