## Supplemental Materials for "Body fat and human cardiovascular ageing"

### Supplemental Material

#### Supplemental Methods

##### Statistical modelling and data standardisation for adiposity phenotypes

For each adiposity phenotype and circulating biomarker, we fitted three linear models: one for males, one for females, and one for both sexes combined.

$$\begin{aligned}\text{Dependent Variable} = \beta_0 + \beta_1 \times \text{Independent Variable 1}\\ + \beta_2 \times \text{Independent Variable 2}\\ + \beta_3 \times \text{Independent Variable 3}\\ + \beta_4 \times \text{Independent Variable 4}\\ + \varepsilon\end{aligned}$$

Additionally, to account for potential effects of body size on adiposity distribution, we adjusted for height-squared to control for variation in stature, following the principles of allometric scaling.[29]

Given that the extracted data included various measurements for adipose tissue, we standardised all available data to ensure accurate statistical calculations. Raw data for visceral adipose tissue (VAT), abdominal subcutaneous adipose tissue (ASAT), muscle adipose tissue infiltration (MATI), liver proton density fat fraction (PDFF), total abdominal adipose tissue (TAAT), android and gynoid adipose tissue mass, total trunk fat mass (TTFM), whole body fat mass (WBFM), apolipoprotein A and B, direct low-density lipoprotein, high-density lipoprotein, triglycerides, and cholesterol underwent Box-Cox normalisation to meet the assumption of normally distributed residuals.

$$y^{(\lambda)} = \begin{cases} \frac{y^{\lambda}-1}{\lambda} & \text{if } \lambda \neq 0, \\ \ln(y) & \text{if } \lambda = 0. \end{cases}$$

When the value of a predictor variable increases by one standard deviation, the outcome variable is expected to increase by the amount of the coefficient associated with that predictor in the regression model. This coefficient indicates the change in the outcome variable for each one standard deviation increase in the predictor variable, while keeping all other variables constant.

$$\beta_{\text{std}} = (\text{SD}_X)\beta/\text{SD}_Y$$

The 95% confidence interval (CI) for the effect estimates was calculated using the formula:

$$\beta \pm 1.96 \times \text{SE}(\beta)$$

The Pearson correlation coefficient was used to examine the linear relationships between clinical variables, such as adipose tissue phenotypes, circulating biomarkers, and their effect on age-delta.

$$r = \frac{\sum(X_i - \overline{X})(Y_i - \overline{Y})}{\sqrt{\sum(X_i - \overline{X})^2 \sum(Y_i - \overline{Y})^2}}$$

##### Quantification of cardiovascular age-delta

We used a pre-trained model developed to predict cardiovascular age using image-derived phenotypes from cardiac MRI, based on 39,559 UK Biobank participants.[9] It was initially trained on 5,065 healthy individuals, free from cardiac, metabolic, or respiratory diseases, with a body mass index below 30. These participants were divided into training (80%, n = 4,019) and test (20%, n = 1,044) sets. CatBoost, a gradient boosting algorithm, was used with default hyperparameters and early stopping rounds set between 50 and 100. Hyperparameter tuning was performed using a 10% validation holdout (n = 403) (using Python package `Optuna`). A further 10% (n = 362) was used for internal early stopping, leaving 3,256 instances for final training. Thirty models with different random seeds were trained, and the one with the lowest mean absolute error on the holdout set was selected. This model predicted age in the remaining 34,147 participants. To correct for bias between predicted age-delta and chronological age, a linear regression was conducted. The regression slope and intercept were used to calculate an offset, which was subtracted from the uncorrected predicted age, yielding the corrected cardiovascular age.

##### Quantile-based categorisation of BMI and corresponding whole body fat mass

To categorise whole body fat mass (WBFM) based on specific Body Mass Index (BMI) quantiles, a systematic approach involving the calculation of empirical cumulative distribution functions (ECDF) for both BMI and WBFM distributions was utilised (Supplementary Figure 1). The detailed steps are as follows:

**Dataset preparation:** The dataset containing BMI and WBFM measurements was loaded and prepared for analysis. Specifically, key quantiles within the BMI distribution were identified and mapped onto the WBFM distribution.

**Empirical cumulative distribution function (ECDF):** For a given dataset $X = \{x_1, x_2, \ldots, x_n\}$, the ECDF is defined as:

$$\hat{F}_n(x) = \frac{1}{n}\sum_{i=1}^{n} \mathbb{1}(x_i \leq x)$$

where $\mathbb{1}(x_i \leq x)$ is the indicator function, which is 1 if $x_i \leq x$ and 0 otherwise. This function provides a stepwise cumulative probability for each value in the dataset.

**Quantile calculation for BMI:** The ranks of the BMI values were calculated, and the ECDF was applied to determine the quantile for each specified BMI value. Specifically, for a BMI value $b$, the quantile $Q_b$ is:

$$Q_b = \min\left(\hat{F}_n(b) \mid b_i \geq b\right)$$

This quantile $Q_b$ indicates the proportion of the dataset with BMI values less than or equal to $b$.

**Mapping BMI quantiles to WBFM:** Using the BMI quantiles obtained, the corresponding WBFM values at these quantiles were identified. For a quantile $Q_b$, the WBFM value $w_b$ is calculated as:

$$w_b = \text{Quantile}(\text{WBFM}, Q_b)$$

where $\text{Quantile}(\text{WBFM}, Q_b)$ is the WBFM value at the specified quantile $Q_b$.

**Visualisation:** The distributions of BMI and WBFM were visualised with specified quantile lines. Histograms were plotted for both distributions, and vertical lines indicating the specified BMI quantiles and corresponding WBFM values were superimposed. This dual-distribution plot provides a clear visual representation of the relationship between BMI and WBFM across the quantiles.

### Supplementary Tables

**Supplementary Table 1. Summary of fat phenotypes by sex and ancestry**. VAT, visceral adipose tissue; ASAT, abdominal subcutaneous adipose tissue; TTFM, total trunk fat mass; WBFM, whole body fat mass; TAAT, total abdominal adipose tissue; MATI, muscle adipose tissue infiltration. IQR, interquartile range.

| Phenotype | Sex | Ancestry | 1st Quartile (0.25) | Median (0.5) | 3rd Quartile (0.75) | IQR |
|---|---|---|---|---|---|---|
| VAT | Female | White background | 1.58 | 2.47 | 3.65 | 2.07 |
| VAT | Female | Black background | 1.50 | 2.33 | 2.92 | 1.42 |
| VAT | Female | Asian background | 1.69 | 2.83 | 3.95 | 2.26 |
| VAT | Female | Mixed background | 1.52 | 2.57 | 3.83 | 2.31 |
| VAT | Male | White background | 3.34 | 4.83 | 6.47 | 3.13 |
| VAT | Male | Black background | 3.18 | 4.67 | 6.45 | 3.26 |
| VAT | Male | Asian background | 3.69 | 4.82 | 6.52 | 2.83 |
| VAT | Male | Mixed background | 3.18 | 5.00 | 6.55 | 3.36 |
| ASAT | Female | White background | 5.82 | 7.75 | 10.15 | 4.33 |
| ASAT | Female | Black background | 6.12 | 8.11 | 9.93 | 3.80 |
| ASAT | Female | Asian background | 5.96 | 8.16 | 11.11 | 5.15 |
| ASAT | Female | Mixed background | 6.17 | 8.03 | 9.87 | 3.70 |
| ASAT | Male | White background | 4.29 | 5.50 | 7.07 | 2.78 |
| ASAT | Male | Black background | 4.42 | 5.91 | 6.73 | 2.31 |
| ASAT | Male | Asian background | 4.69 | 5.84 | 7.11 | 2.41 |
| ASAT | Male | Mixed background | 4.07 | 5.15 | 6.59 | 2.52 |
| TTFM | Female | White background | 9.90 | 12.80 | 16.20 | 6.30 |
| TTFM | Female | Black background | 10.70 | 13.20 | 17.00 | 6.30 |
| TTFM | Female | Asian background | 10.25 | 13.00 | 17.85 | 7.60 |
| TTFM | Female | Mixed background | 9.85 | 12.40 | 16.45 | 6.60 |
| TTFM | Male | White background | 10.80 | 13.50 | 16.60 | 5.80 |
| TTFM | Male | Black background | 10.97 | 13.70 | 17.25 | 6.28 |
| TTFM | Male | Asian background | 11.70 | 14.20 | 16.65 | 4.95 |
| TTFM | Male | Mixed background | 10.75 | 12.70 | 16.05 | 5.30 |
| WBFM | Female | White background | 19.80 | 25.00 | 31.40 | 11.60 |
| WBFM | Female | Black background | 21.30 | 26.30 | 32.00 | 10.70 |
| WBFM | Female | Asian background | 20.18 | 25.95 | 33.90 | 13.72 |
| WBFM | Female | Mixed background | 20.30 | 24.45 | 32.12 | 11.82 |
| WBFM | Male | White background | 17.00 | 21.10 | 26.30 | 9.30 |
| WBFM | Male | Black background | 17.43 | 21.65 | 27.23 | 9.80 |
| WBFM | Male | Asian background | 18.65 | 22.20 | 26.75 | 8.10 |
| WBFM | Male | Mixed background | 16.82 | 20.55 | 25.43 | 8.60 |
| TAAT | Female | White background | 0.49 | 0.56 | 0.62 | 0.14 |
| TAAT | Female | Black background | 0.48 | 0.55 | 0.61 | 0.12 |
| TAAT | Female | Asian background | 0.48 | 0.57 | 0.65 | 0.17 |
| TAAT | Female | Mixed background | 0.50 | 0.56 | 0.63 | 0.13 |
| TAAT | Male | White background | 0.40 | 0.46 | 0.52 | 0.13 |
| TAAT | Male | Black background | 0.39 | 0.45 | 0.52 | 0.13 |
| TAAT | Male | Asian background | 0.41 | 0.46 | 0.52 | 0.11 |
| TAAT | Male | Mixed background | 0.37 | 0.45 | 0.52 | 0.15 |
| MATI | Female | White background | 6.63 | 7.66 | 8.91 | 2.28 |
| MATI | Female | Black background | 6.58 | 7.67 | 8.61 | 2.03 |
| MATI | Female | Asian background | 6.77 | 7.91 | 9.22 | 2.45 |
| MATI | Female | Mixed background | 6.88 | 7.96 | 8.93 | 2.05 |
| MATI | Male | White background | 5.66 | 6.58 | 7.72 | 2.06 |
| MATI | Male | Black background | 5.53 | 6.84 | 7.58 | 2.05 |
| MATI | Male | Asian background | 5.67 | 6.73 | 7.76 | 2.09 |
| MATI | Male | Mixed background | 5.59 | 6.44 | 7.45 | 1.86 |

**Supplementary Table 2. Genetic instruments for Mendelian randomisation**. Gluteofemoral adipose tissue (GFAT), visceral adipose tissue (VAT), abdominal subcutaneous adipose tissue (ASAT) genetic variants Used in MR analysis on causal association to the cardiovascular age (outcome). Columns include the dbSNP ID, GRCh37 position, effect allele and the effect allele frequency (EAF), and the Genome-Wide Association Study (GWAS) stats for each Single Nucleotide Polymorphism (SNP) on exposure and outcome (Beta, SE, *P* value).

| | | | | | | | | Exposure | | | Outcome | |
|---|---|---|---|---|---|---|---|---|---|---|---|---|
| | **SNP** | **CHROM** | **POS** | Effect Allele | Other Allele | EAF (Exposure) | Beta | SE | *P* value | Beta | SE | *P* value |
| Exposure: GFAT (ID: gfatadjbmi3) | | | | | | | | | | | | |
| 1 | rs10044492 | 5 | 167634 | C | T | 0.732286 | -0.0478462 | 0.00805882 | 5.30E-09 | 0.0371827 | 0.0714443 | 0.602758 |
| 2 | rs10501153 | 11 | 563572 | C | T | 0.677439 | -0.0439302 | 0.0076186 | 5.90E-09 | -0.071416 | 0.0675313 | 0.290281 |
| 3 | rs11205303 | 1 | 165637 | T | C | 0.596291 | -0.0392991 | 0.00724463 | 1.70E-08 | 0.126362 | 0.064481 | 0.0500429 |
| 4 | rs114078082 | 6 | 544713 | G | A | 0.958096 | 0.138123 | 0.0177313 | 2.00E-15 | -0.126356 | 0.154574 | 0.41368 |
| 5 | rs12814794 | 12 | 460442 | G | A | 0.247599 | -0.0721238 | 0.00826435 | 1.60E-18 | -0.118934 | 0.073571 | 0.105977 |
| 6 | rs13099700 | 3 | 226439 | A | G | 0.721677 | 0.0471756 | 0.00794479 | 7.90E-09 | -0.055496 | 0.0705424 | 0.431461 |
| 7 | rs13142096 | 4 | 730465 | A | G | 0.727366 | -0.0470762 | 0.00801245 | 8.40E-09 | 0.160797 | 0.0712461 | 0.0240202 |
| 8 | rs13389219 | 2 | 18149 | C | T | 0.606906 | -0.0731577 | 0.00725328 | 3.00E-23 | 0.0865677 | 0.0644427 | 0.179176 |
| 9 | rs1469246 | 4 | 166086 | G | A | 0.671272 | -0.0540125 | 0.0075806 | 9.40E-13 | 0.104254 | 0.0673262 | 0.121513 |
| 10 | rs1907218 | 10 | 152883 | T | C | 0.314461 | -0.0488068 | 0.00764797 | 3.60E-10 | 0.00286916 | 0.0677417 | 0.966216 |
| 11 | rs2082162 | 5 | 598391 | G | T | 0.412166 | -0.038786 | 0.00725607 | 2.20E-08 | -0.0525403 | 0.0643704 | 0.414382 |
| 12 | rs2267373 | 22 | 471412 | C | T | 0.418883 | 0.0461893 | 0.00722912 | 1.40E-10 | -0.00671632 | 0.0641773 | 0.916652 |
| 13 | rs2300669 | 3 | 620155 | C | A | 0.615258 | -0.0421979 | 0.00727957 | 4.40E-09 | -0.0163743 | 0.0645603 | 0.799784 |
| 14 | rs2943653 | 2 | 24424 | C | T | 0.325957 | 0.0751004 | 0.0075487 | 6.90E-23 | 0.0105112 | 0.0671747 | 0.875659 |
| 15 | rs2955617 | 17 | 196865 | C | A | 0.348318 | -0.0418669 | 0.00747226 | 1.20E-08 | 0.0837415 | 0.066523 | 0.2081 |
| 16 | rs3822072 | 4 | 104928 | G | A | 0.545788 | 0.0478524 | 0.00712951 | 4.90E-12 | -0.145465 | 0.0631734 | 0.021307 |
| 17 | rs3936511 | 5 | 681508 | A | G | 0.808734 | 0.0815883 | 0.00901579 | 3.90E-20 | -0.0777822 | 0.0805452 | 0.334204 |
| 18 | rs4450871 | 4 | 879764 | A | G | 0.554794 | -0.0383337 | 0.0071357 | 3.10E-08 | 0.0306861 | 0.0632487 | 0.627562 |
| 19 | rs4759309 | 12 | 682774 | G | A | 0.221429 | -0.0443202 | 0.00853853 | 4.20E-08 | -0.00480906 | 0.0763794 | 0.949797 |
| 20 | rs546560809 | 4 | 134628 | T | G | 0.961183 | 0.0981811 | 0.0184826 | 2.50E-08 | -0.237203 | 0.1644 | 0.149074 |
| 21 | rs71304101 | 3 | 297793 | G | A | 0.879021 | -0.0618006 | 0.0108921 | 1.70E-09 | -0.0166699 | 0.0969471 | 0.86348 |
| 22 | rs7133378 | 12 | 14789 | G | A | 0.679988 | -0.0876372 | 0.00761231 | 5.60E-29 | 0.245266 | 0.0674336 | 0.000276149 |
| 23 | rs72959041 | 6 | 132816 | G | A | 0.952589 | 0.195251 | 0.0168711 | 3.20E-32 | 0.0319367 | 0.148534 | 0.829759 |
| 24 | rs8075019 | 17 | 444844 | G | A | 0.872483 | 0.0634833 | 0.0107754 | 2.30E-10 | -0.0383858 | 0.0965028 | 0.690803 |
| 25 | rs998584 | 6 | 677005 | C | A | 0.516864 | 0.07952 | 0.00711591 | 6.10E-31 | -0.188181 | 0.0631683 | 0.00289384 |
| Exposure: VAT (ID: vatadjbmi3) | | | | | | | | | | | | |
| 1 | rs11031796 | 11 | 498806 | G | A | 0.611683 | 0.0524669 | 0.00728182 | 5.10E-14 | -0.0451491 | 0.0646797 | 0.485157 |
| 2 | rs11992444 | 8 | 565812 | G | T | 0.491592 | -0.0779981 | 0.00709912 | 1.30E-29 | -0.10997 | 0.0632918 | 0.0823087 |
| 3 | rs12089366 | 1 | 225654 | C | T | 0.776758 | 0.0580131 | 0.00855551 | 9.40E-12 | -0.0365977 | 0.0765896 | 0.632767 |
| 4 | rs1329254 | 10 | 50472 | C | T | 0.369978 | 0.0418348 | 0.00733732 | 1.40E-08 | -0.081558 | 0.0651416 | 0.210576 |
| 5 | rs1635851 | 7 | 463016 | C | T | 0.414446 | 0.040899 | 0.00720247 | 3.80E-08 | -0.0305015 | 0.064236 | 0.634908 |
| 6 | rs30351 | 5 | 680081 | G | A | 0.26445 | 0.0705595 | 0.00808072 | 1.10E-16 | -0.033718 | 0.0724539 | 0.641669 |
| 7 | rs35932591 | 2 | 142074 | C | T | 0.878816 | 0.0606447 | 0.0107886 | 3.80E-08 | 0.0379672 | 0.0967951 | 0.694882 |
| 8 | rs3731861 | 2 | 233585 | T | C | 0.62196 | -0.0382281 | 0.00730702 | 4.70E-08 | 0.0661645 | 0.0654018 | 0.311708 |
| 9 | rs4307676 | 11 | 66432 | G | A | 0.833929 | 0.0535154 | 0.00949868 | 8.80E-09 | 0.104528 | 0.0840798 | 0.213805 |
| 10 | rs4872393 | 8 | 575554 | G | A | 0.7734 | -0.0601652 | 0.00844375 | 2.00E-12 | 0.286302 | 0.0758622 | 0.00016098 |
| 11 | rs56006999 | 1 | 224281 | C | T | 0.821332 | 0.0536419 | 0.00922851 | 3.60E-09 | 0.0485811 | 0.0830658 | 0.558653 |
| 12 | rs56082403 | 3 | 171233 | T | C | 0.593073 | -0.0556812 | 0.00723565 | 6.90E-14 | 0.0200518 | 0.0647187 | 0.756692 |
| 13 | rs577721086 | 6 | 132815 | T | C | 0.952316 | -0.117892 | 0.01673 | 5.20E-13 | 0.060537 | 0.147915 | 0.682346 |
| 14 | rs7133378 | 12 | 14789 | G | A | 0.679988 | 0.0458609 | 0.00758948 | 6.60E-10 | 0.245266 | 0.0674336 | 0.000276149 |
| 15 | rs72810972 | 5 | 192967 | G | T | 0.716377 | -0.0539032 | 0.00784907 | 2.30E-12 | -0.0534721 | 0.0694006 | 0.441019 |
| 16 | rs7933253 | 11 | 762768 | T | C | 0.0482717 | 0.0978848 | 0.0167983 | 1.30E-08 | 0.249213 | 0.152765 | 0.102826 |
| 17 | rs998584 | 6 | 677005 | C | A | 0.516864 | -0.0570321 | 0.00708399 | 1.80E-15 | -0.188181 | 0.0631683 | 0.00289384 |
| Exposure: ASAT (ID: asatadjbmi3) | | | | | | | | | | | | |
| 1 | rs1159619 | 6 | 132699 | C | A | 0.544561 | 0.0457682 | 0.0071716 | 1.20E-10 | 0.0367581 | 0.0630816 | 0.560095 |
| 2 | rs11709077 | 3 | 297621 | G | A | 0.88011 | -0.0696508 | 0.0110061 | 1.70E-10 | -0.00247357 | 0.0973027 | 0.979719 |
| 3 | rs13322435 | 3 | 171233 | A | G | 0.59074 | 0.0570714 | 0.00731172 | 2.40E-15 | 0.028349 | 0.0647402 | 0.66147 |
| 4 | rs17205757 | 15 | 860493 | A | G | 0.673992 | -0.0415864 | 0.00765851 | 3.20E-08 | 0.00483059 | 0.0676445 | 0.943071 |
| 5 | rs1779445 | 1 | 150977 | T | C | 0.194296 | -0.0493135 | 0.00906223 | 1.90E-08 | 0.101266 | 0.0806485 | 0.209254 |
| 6 | rs1815172 | 15 | 108655 | C | T | 0.476133 | -0.0594399 | 0.00716879 | 6.80E-17 | -0.0705533 | 0.0630489 | 0.263139 |
| 7 | rs2302209 | 19 | 448214 | C | T | 0.7193 | -0.0461843 | 0.00799643 | 3.40E-09 | -0.114164 | 0.0703054 | 0.104423 |
| 8 | rs2943647 | 2 | 244257 | T | C | 0.348169 | 0.0432106 | 0.00746638 | 5.80E-09 | 0.016165 | 0.0658683 | 0.806138 |
| 9 | rs3850625 | 1 | 217777 | G | A | 0.88219 | -0.0786112 | 0.0110023 | 1.80E-12 | -0.0327814 | 0.0976972 | 0.737219 |
| 10 | rs3936510 | 5 | 681509 | G | T | 0.798132 | -0.0629779 | 0.00887846 | 5.00E-13 | -0.107168 | 0.0790466 | 0.175189 |
| 11 | rs4731702 | 7 | 140174 | C | T | 0.512779 | -0.0471297 | 0.00716051 | 9.20E-11 | 0.0660724 | 0.0632043 | 0.295856 |
| 12 | rs55744247 | 5 | 641338 | G | A | 0.795882 | -0.0534533 | 0.0088675 | 5.10E-10 | -0.113855 | 0.0776773 | 0.142731 |
| 13 | rs7538503 | 1 | 243742 | A | G | 0.710369 | -0.0474118 | 0.00787047 | 8.40E-10 | 0.00713499 | 0.0695653 | 0.918309 |
| 14 | rs8077609 | 17 | 190864 | A | C | 0.673982 | 0.0420664 | 0.00765749 | 1.10E-08 | 0.0845527 | 0.0676265 | 0.211204 |

**Supplementary Table 3. Mendelian randomisation**. Results for the association between body mass index (BMI), height adjusted gluteofemoral adipose tissue (GFAT), visceral adipose tissue (VAT), abdominal subcutaneous adipose tissue (ASAT) and Cardiovascular age-delta.

| **Outcome** | **Exposure** | **Method** | **Number of SNPs** | **Beta** | **Standard Error** | ***P* value** |
|---|---|---|---|---|---|---|
| Cardiovascular age-delta | GFAT adjusted for BMI | MR Egger | 25 | -0.9174 | 0.8089 | 0.2684 |
| Cardiovascular age-delta | GFAT adjusted for BMI | Weighted Median | 25 | -0.9183 | 0.3809 | 0.0159 |
| Cardiovascular age-delta | GFAT adjusted for BMI | Inverse Variance Weighted | 25 | -0.9576 | 0.2877 | 0.0009 |
| Cardiovascular age-delta | GFAT adjusted for BMI | Simple Mode | 25 | -0.4396 | 0.8312 | 0.6017 |
| Cardiovascular age-delta | GFAT adjusted for BMI | Weighted Mode | 25 | -0.3853 | 0.8455 | 0.6527 |
| Cardiovascular age-delta | VAT adjusted for BMI | MR Egger | 17 | 1.1595 | 2.2425 | 0.6126 |
| Cardiovascular age-delta | VAT adjusted for BMI | Weighted Median | 17 | -0.1102 | 0.4951 | 0.8238 |
| Cardiovascular age-delta | VAT adjusted for BMI | Inverse Variance Weighted | 17 | 0.3566 | 0.5343 | 0.5044 |
| Cardiovascular age-delta | VAT adjusted for BMI | Simple Mode | 17 | -0.3572 | 0.8315 | 0.6733 |
| Cardiovascular age-delta | VAT adjusted for BMI | Weighted Mode | 17 | -0.1982 | 0.8011 | 0.8078 |
| Cardiovascular age-delta | ASAT adjusted for BMI | MR Egger | 14 | 1.1547 | 2.0570 | 0.5849 |
| Cardiovascular age-delta | ASAT adjusted for BMI | Weighted Median | 14 | 0.4908 | 0.4533 | 0.2789 |
| Cardiovascular age-delta | ASAT adjusted for BMI | Inverse Variance Weighted | 14 | 0.6128 | 0.3656 | 0.0937 |
| Cardiovascular age-delta | ASAT adjusted for BMI | Simple Mode | 14 | 0.3370 | 0.8015 | 0.6810 |
| Cardiovascular age-delta | ASAT adjusted for BMI | Weighted Mode | 14 | 0.4573 | 0.7401 | 0.5473 |

### Supplementary Figures

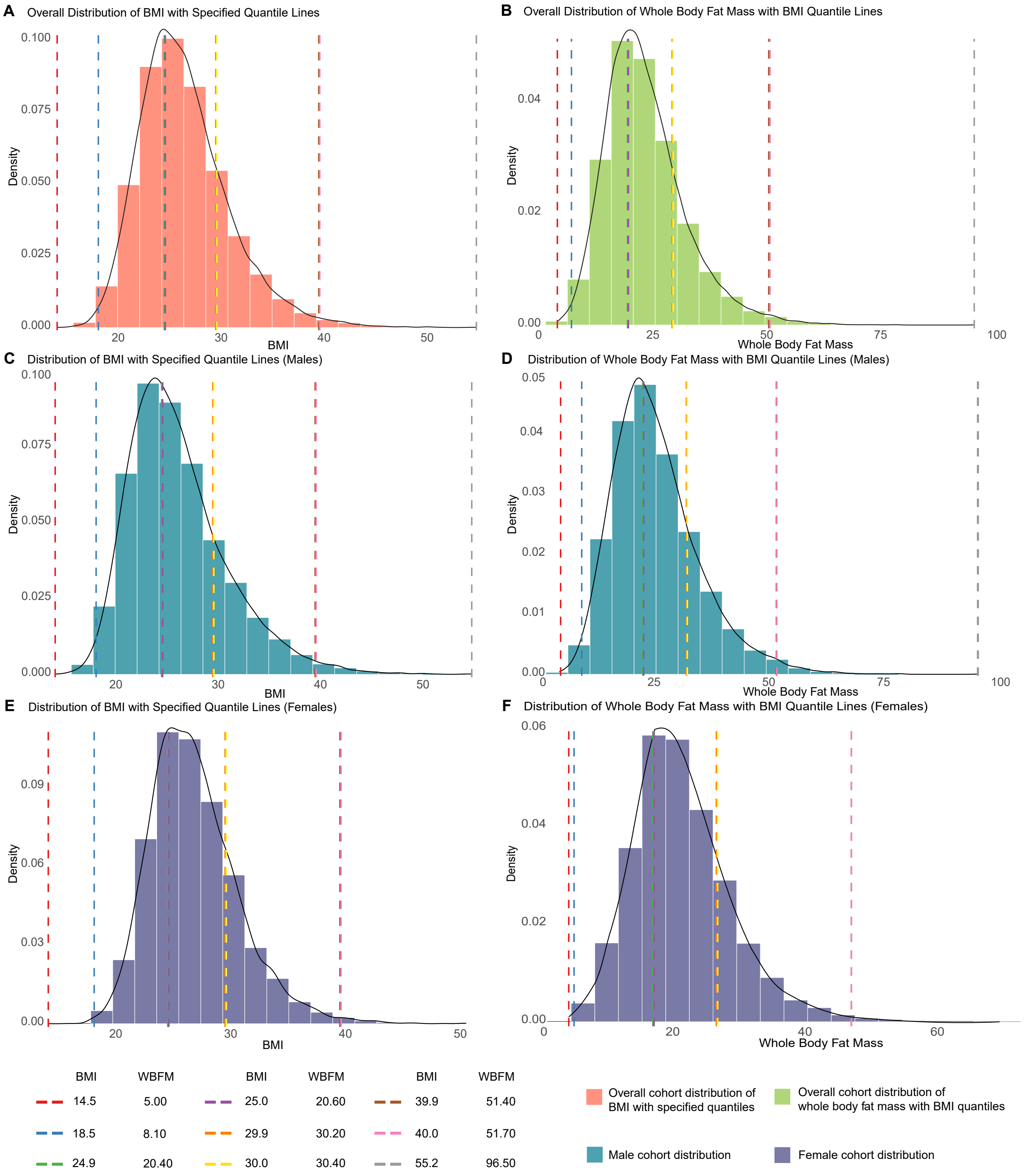


**Supplementary Figure 1. Categorisation approach for whole body fat mass based on BMI quantiles.** Density distributions of body mass index (BMI) and whole body fat mass (WBFM) for different cohorts with specified quantile lines; Panel **A** depicts the overall BMI distribution with quantile lines, showing the spread of BMI values in the entire cohort; Panel **B** illustrates the WBFM distribution for the entire cohort with BMI quantile lines; Panel **C** displays the BMI distribution for males with marked quantiles; Panel **D** represents the WBFM distribution for males, annotated with BMI quantiles; Panel **E** shows the BMI distribution for females similarly; Panel **F** demonstrates the WBFM distribution for females, indicating the density and quantile lines based on BMI. Quantile markers are provided for key percentiles, highlighting the central tendencies and distribution spread within each cohort.

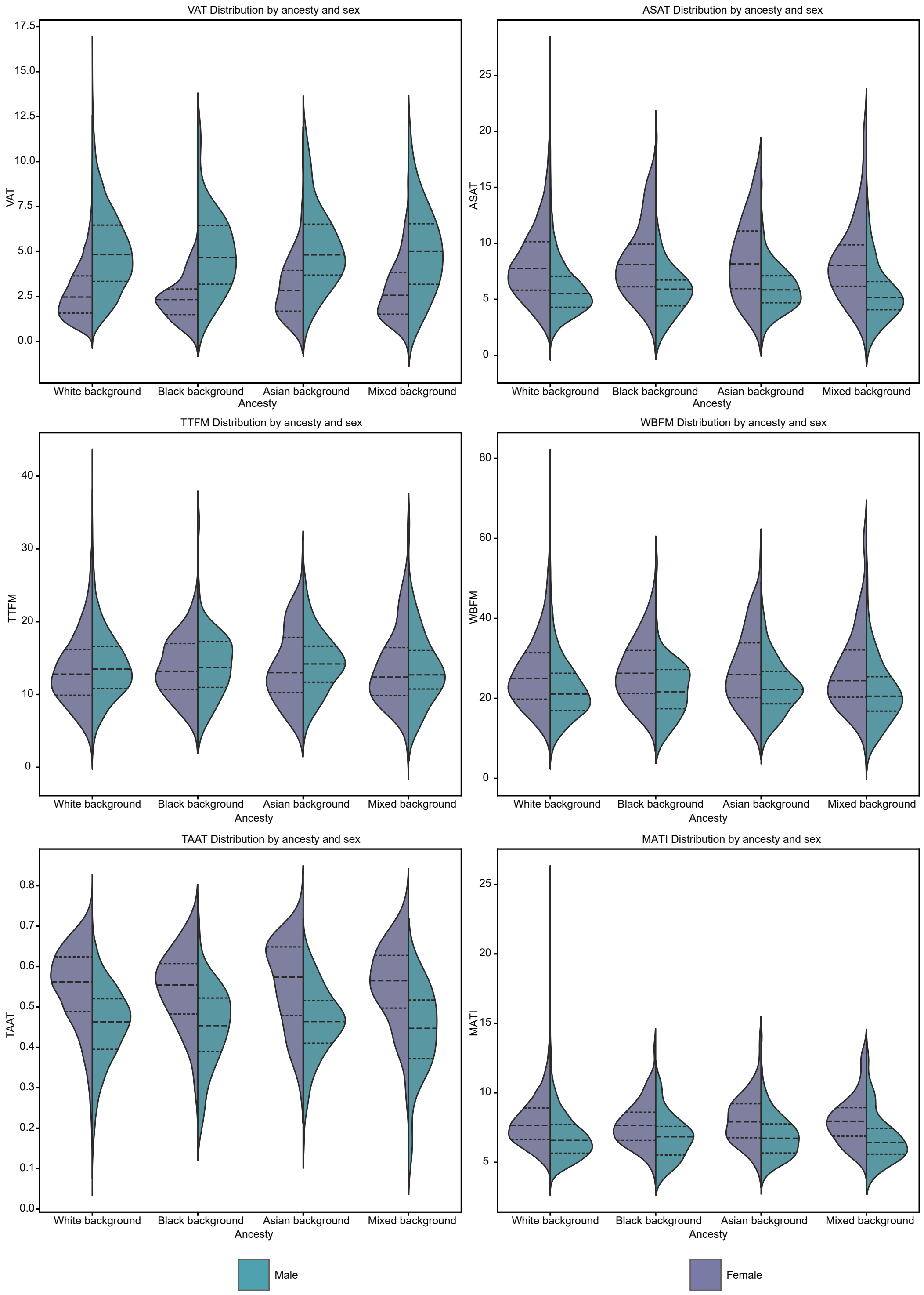


**Supplementary Figure 2. Adiposity distributions by ancestry**. Violin plots show the distribution of fat phenotypes stratified by ancestry and sex: White background (n=20631, 97.12%), ; Black background (n=128, 0.60%); Asian background (n=262, 1.24%); Mixed background (n=220, 1.04%). Dashed lines show each quartile and values are shown in Supplementary Table 1. VAT, visceral adipose tissue; ASAT, abdominal subcutaneous adipose tissue; MATI, muscle adipose tissue infiltration; TTFM, total trunk fat mass; WBFM, whole body fat mass; TAAT, total abdominal adipose tissue.

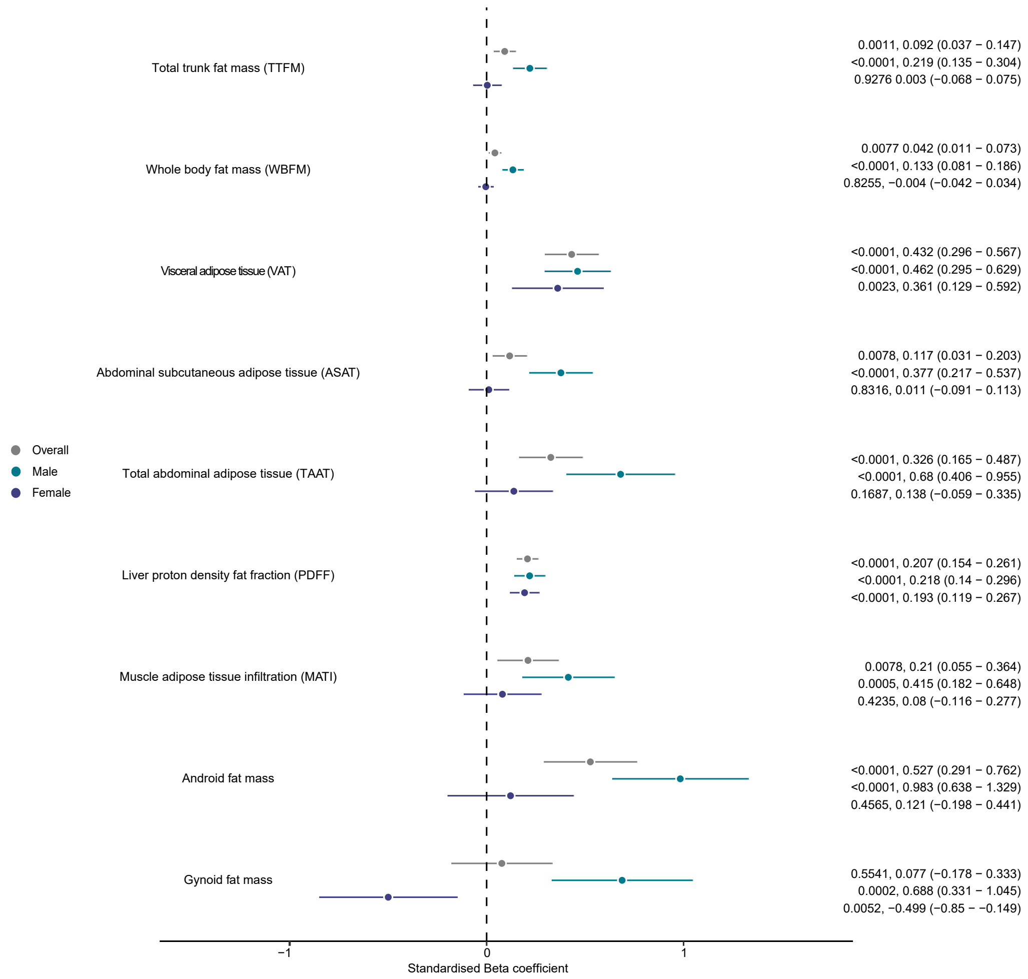


**Supplementary Figure 3. Height$^2$-adjusted effect of adiposity phenotypes on cardiovascular age-delta**. Linear regression analysis of quantitative adipose tissue traits (n=21,241 of which 5,168 had android and gynoid fat mass values) with height$^2$-adjusted cardiovascular age-delta as the dependent variable. *P* values, standardised beta-coefficient point estimates, and 95% confidence intervals are shown, stratified by sex. The adipose tissue traits include gynoid fat mass, android fat mass, muscle adipose tissue infiltration (MATI), liver proton density fat fraction (PDFF), total abdominal adipose tissue (TAAT), abdominal subcutaneous adipose tissue (ASAT), visceral adipose tissue (VAT), whole body fat mass (WBFM), and total trunk fat mass (TTFM).

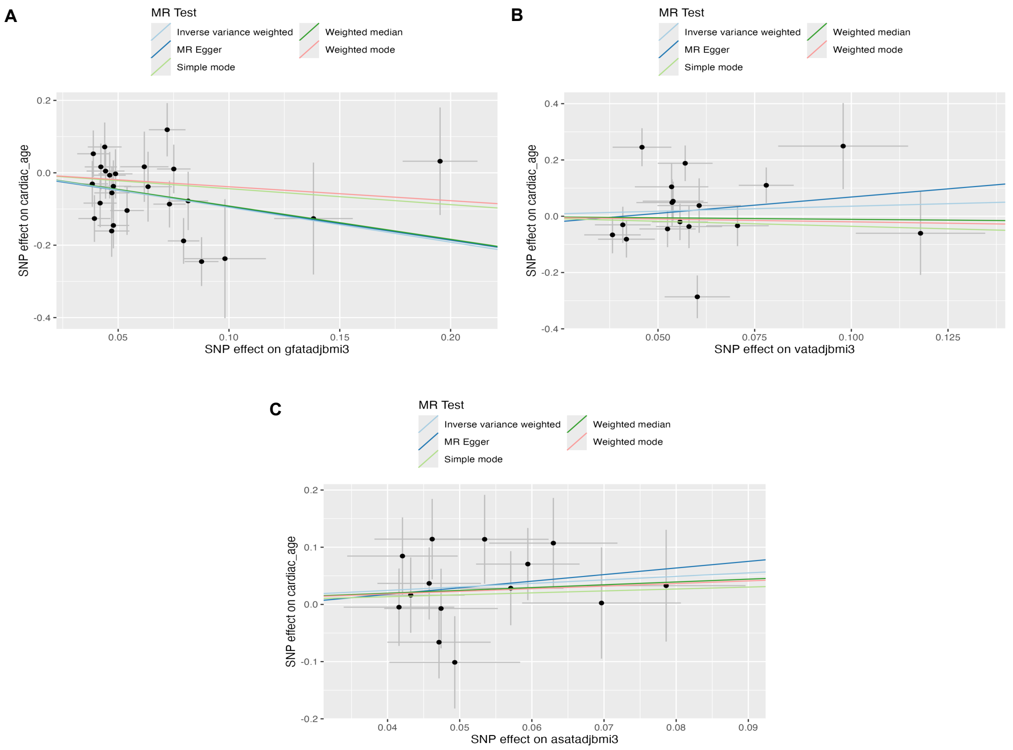


**Supplementary Figure 4. Single nucleotide polymorphism (SNP) effects of body fat on cardiovascular age**. Mendelian randomisation (MR) analysis of BMI adjusted **A** gluteofemoral adipose tissue, **B** visceral adipose tissue, and **C** abdominal subcutaneous adipose tissue as exposure, with cardiovascular age as outcome. Genetic instruments for body fat were selected from a published GWAS.[27] The effects ($\beta$) of the exposure variable-increasing allele at independent SNPs ($r^2 < 0.001$) reaching $P < 5e\text{-}8$ are plotted as datapoints and associated standard errors are represented as lines extending from datapoints. The plots were produced using the R package `TwoSampleMR`. See Supplementary Table 2 for full MR results and Supplementary Table 3 for full list of SNP IDs.
